# Determinants and clinical implications of discharge timing after catheter ablation for atrial tachycardia

**DOI:** 10.64898/2026.02.07.26345799

**Authors:** Ann-Kathrin Kahle, Florian Doldi, Piotr Foszcz, Omar Anwar, Melanie A. Gunawardene, Annika Haas, Fares-Alexander Alken, Katharina Scherschel, Jasper Junker, Julia Mehrhoff, Karim Abudaher, Armin Luik, Andreas Metzner, Paulus Kirchhof, Arian Sultan, Stephan Willems, Lars Eckardt, Ernan Zhu, Christian Meyer

## Abstract

**Aims:** Early discharge after electrophysiological procedures has gained increasing attention. However, definition of patient- and procedure-related prerequisites for successful and safe discharge strategies after atrial tachycardia (AT) ablation remains unknown. We therefore evaluated patient characteristics, procedural features, and outcomes according to index length of stay (LOS) following AT ablation.

**Methods and results:** The multicenter observational SATELLITE registry enrolled consecutive patients undergoing AT rhythm control. Patients were stratified by LOS (≤1, 2 and >2 nights) after catheter ablation. Among 670 patients (67 [IQR 56–75] years, 54.9% male), LOS was ≤1 night in 13.9%, 2 nights in 41.9% and >2 nights in 44.2%. LOS was only modestly predictable from clinical characteristics including age, sex, atrial fibrillation and prior atrial ablation (AUC 0.73). Discrimination improved after inclusion of procedural variables and early post-procedural events (AUC 0.77; *P*=0.0300), consistent with an increase in left atrial procedures (26.5% vs. 76.0% vs. 80.8%; *P*<0.0001), acute minor complications (3.2% vs. 2.5% vs. 14.5%; *P*<0.0001) and early recurrences of atrial arrhythmia (2.2% vs. 6.8% vs. 21.3%; *P*<0.0001). During 2.8±3.0 years of follow-up, LOS did not predict long-term outcomes including subsequent cardiovascular hospitalization (HR 1.19, 95% CI 0.78–1.81; *P*=0.4175).

**Conclusion:** Despite multiple comorbidities, most patients undergoing AT ablation need up to 2 nights of hospitalization. However, prolonged hospital stays before successful and safe discharge are common and associated with acute minor complications and early recurrences of atrial arrhythmia rather than comorbidities. Accordingly, discharge timing largely reflects the immediate peri-procedural clinical course, therefore challenging purely logistics-driven planning.

**Key Learning Points:** *What is already known:*

- Early discharge after electrophysiological procedures has gained increasing attention.
- Definition of patient- and procedure-related prerequisites for successful and safe discharge strategies after atrial tachycardia (AT) ablation remains unknown.

*What this study adds:*

- Despite multiple comorbidities, most patients undergoing AT ablation need up to 2 nights of hospitalization.
- Prolonged hospital stays before successful and safe discharge are common and associated with acute minor complications and early recurrences of atrial arrhythmia rather than comorbidities.
- Discharge timing largely reflects the immediate peri-procedural clinical course, therefore challenging purely logistics-driven planning

*Structured Graphical Abstract:*

- Despite multiple comorbidities, most patients undergoing AT ablation need up to 2 nights of hospitalization. However, prolonged hospital stays before successful and safe discharge are common and associated with acute minor complications and early recurrences of atrial arrhythmia rather than comorbidities. Accordingly, discharge timing largely reflects the immediate peri-procedural clinical course, therefore challenging purely logistics-driven planning. 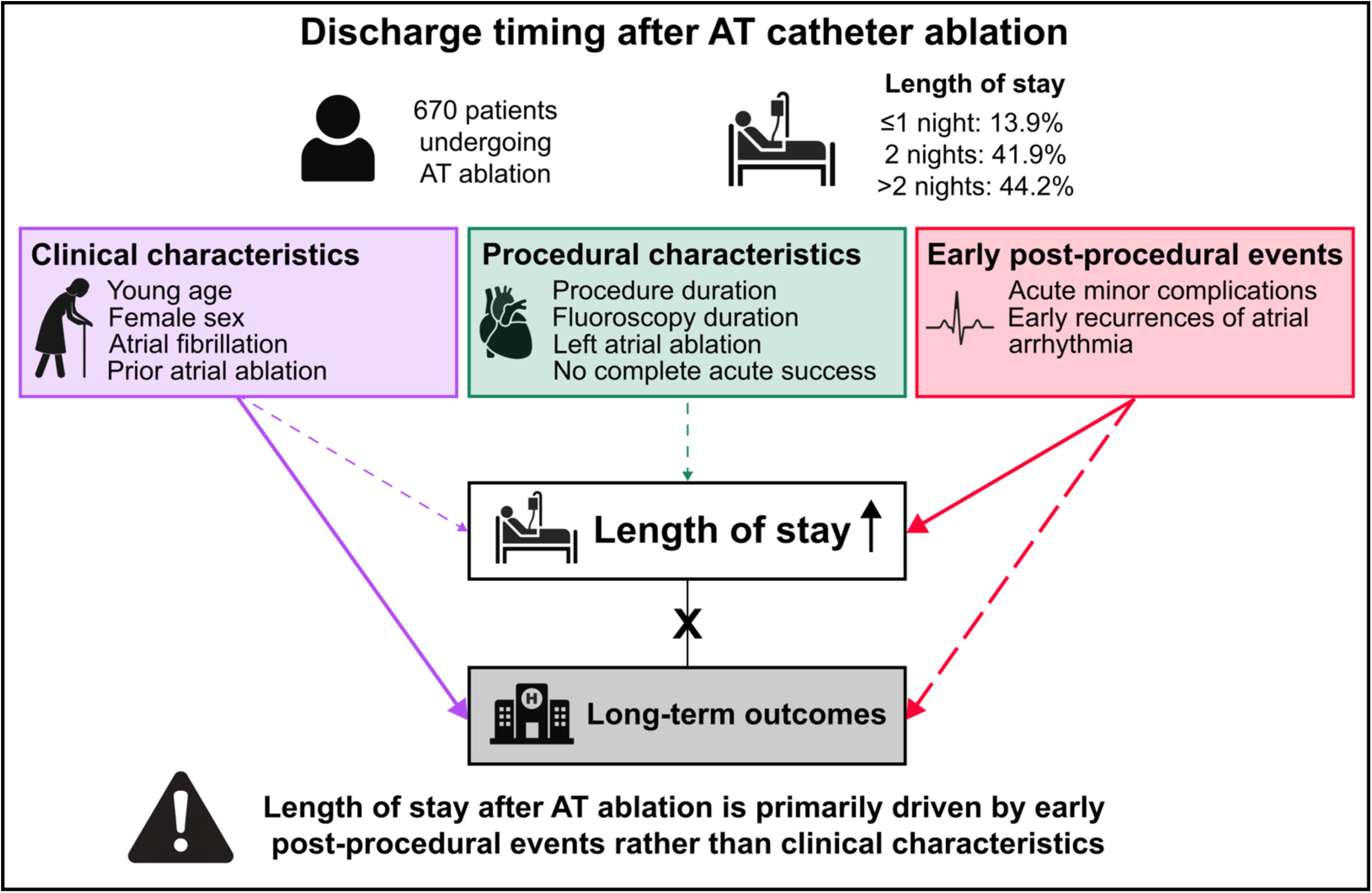

## Introduction

Atrial tachycardias (AT) are increasingly encountered in clinical practice and are associated with substantial symptom burden and recurrent healthcare utilization.^1^ Catheter ablation represents the most effective rhythm control strategy for regular AT,^2^ yet patients frequently present with multiple comorbidities and demanding atrial substrates eventually resulting in complex procedures and requiring structured post-procedural monitoring.^3^ Meanwhile, early discharge strategies have gained attention across electrophysiological procedures, driven by advances in procedural safety, catheter technologies, peri-procedural care pathways and economic pressure.^4,5^ As demonstrated in a recent meta-analysis conducted by the European Heart Rhythm Association Health Economics Committee, same-day discharge may be feasible in selected atrial fibrillation (AF) ablation cohorts without an increase in post-discharge unplanned medical contacts or rehospitalization compared with overnight stay.^6^ However, its applicability to AT ablation is uncertain, given distinct arrhythmia mechanisms, procedural complexity, and patient profiles. In particular, definition of patient- and procedure-related prerequisites for successful and safe discharge strategies after AT ablation remains unknown. Against this background, the present study systematically evaluated patient characteristics, procedural features, early post-procedural events, and long-term outcomes according to index length of stay (LOS) following AT ablation in a large multicenter cohort.

## Methods

### Study population

The multicenter observational SATELLITE (Systematic Assessment of Treatment Effectiveness for Long-Term Management of Stable Atrial Tachycardia in Inpatient and Outpatient Environments) registry enrolled consecutive patients undergoing AT rhythm control in 5 tertiary arrhythmia centers until 2025 (NCT07239804). In the present analysis, patients were stratified by index LOS following AT ablation. LOS was analyzed as a pragmatic endpoint reflecting both medical needs and clinical workflow decisions. Given the potential influence of diagnosis-related group structures and organizational factors, LOS analyses were complemented by acute minor complications and early recurrences of atrial arrhythmia to capture medically driven in-hospital pathways.

### Electrophysiological study

Mapping and catheter ablation were conducted under deep sedation, as previously described.^7^ A steerable 6-F decapolar diagnostic catheter (Inquiry^TM^, 2-5-2 mm spacing, St. Jude Medical, St. Paul, MN; or Dynamic DECA / EP XT, Boston Scientific, Marlborough, MA) was placed in the coronary sinus as a reference, and an open-irrigated 3.5-mm tip catheter (Thermocool®, D- or F-Type-Curve; Biosense Webster, CA; or IntellaNav MiFi OI®; Boston Scientific; or IntellaNav Stablepoint^TM^; Boston Scientific) was used for ablation. Electroanatomical mapping was performed with a multielectrode catheter (Orion^TM^; Boston Scientific or Pentaray; Biosense Webster or Lasso catheter; Biosense Webster). The left atrium (LA) was accessed by double transseptal access after single or double transseptal puncture using a fixed curve long sheath (SL0 or SL1, 8-F; St. Jude Medical, HeartSpan® 55°, Merit Medical). Heparin was administered intravenously to maintain an activated clotting time >300 s in LA procedures.

For patients in sinus rhythm at the beginning of the procedure, programmed stimulation or burst atrial pacing was performed.^8^ The clinical AT was assumed when cycle length and P wave morphology matched the preprocedural 12-channel electrocardiogram documentation.

### Electroanatomical mapping and catheter ablation

Activation and voltage mapping was performed for inducible or ongoing AT.^8^ Maps were considered complete when the entire chamber anatomy was reconstructed with the best achievable electrode-tissue contact. Activation maps were created under standard automatic beat acceptance criteria. Wavefront propagation, activation patterns, areas of slow conduction, anatomical and functional barriers, and lines of block were reviewed to target the critical isthmus or site of earliest activation.^9^ Additional entrainment mapping was performed at the operator’s discretion.

Radiofrequency current was applied using a maximum power of 40 W and peak tip temperature of 42°C. For macro-reentrant tachycardia, the critical isthmus was targeted in order to transect the circuit by linear ablation. Localized reentrant AT were targeted at their critical site with creation of linear lesions bridging the nearest anatomical-surgical barriers, focal AT by ablation at the site of earliest activation. Endpoints of previously performed ablations such as pulmonary vein isolation or integrity of lines were verified and completed, if appropriate. Bidirectional block was confirmed by differential pacing and/or repeat mapping, whenever linear lesions were generated. Acute, complete procedural success was defined as termination of all stable, inducible AT and non-inducibility of further sustained tachycardias confirmed by routine atrial burst pacing or programmed stimulation.^10^

### Follow-up

All patients were routinely followed-up at 3 months after the index procedure, with subsequent follow-ups every 6 to 12 months.^10^ Pacemaker and implantable cardioverter defibrillators were interrogated through remote monitoring. 12-channel as well as 24-hour Holter electrocardiograms were performed in all patients without implantable devices for further assessment. Arrhythmia recurrences were defined as episodes lasting >30 s and occurring >2 months after ablation, early recurrences of atrial arrhythmia as occurring <2 months after ablation.^11,12^ In the case of a recurrent, sustained or symptomatic arrhythmia, patients were considered for a repeat ablation.^12,13^

### Statistical analysis

Continuous variables are presented as mean ± SD or median (interquartile range: 25^th^ – 75^th^ percentile) and were compared by one-way ANOVA or Kruskal–Wallis test including multiple comparisons. Categorical variables are presented as counts (percentage) and were compared by chi-square test.

Predictability of LOS was assessed using hierarchical logistic regression models with prolonged hospitalization defined as a binary outcome. Models were sequentially constructed by first incorporating baseline clinical characteristics, followed by the addition of procedural variables and early post-procedural events. Discriminative performance was quantified using the area under the receiver operating characteristic curve (AUC), and incremental differences between nested models were formally compared using DeLong’s test. For risk stratification, patients were categorized into tertiles of the linear predictor (low, intermediate, high clinical LOS risk), and observed proportions of LOS ≥2 nights were compared across strata. Model calibration was assessed graphically by plotting observed vs. mean predicted risk across deciles of the predicted probability of LOS ≥2 nights.

Logistic regression analyses were conducted to identify predictors of acute minor complications and early recurrences of atrial arrhythmia as well as the composite of both. Given the low event rates, bias-reduced logistic regression using Firth’s penalized likelihood was applied to obtain stable effect estimates and mitigate small-sample bias.

Event-free survival was estimated using the Kaplan-Meier method with the log-rank test. Because LOS was influenced by early post-procedural events and subject to immortal time and reverse causation, conventional Cox models were complemented by a landmark analysis at day 2 restricted to clinically stable patients to assess whether an additional night of hospitalization provides prognostic benefit.^14^ Patients with acute complications or early recurrences of atrial arrhythmia were excluded. Analyses were restricted to those discharged after ≤1 or 2 nights. Time-to-event analyses for long-term outcomes were performed using multivariable Cox proportional hazards models, with the landmark set as time zero. Models were adjusted for age, sex, CHA₂DS₂-VA score, AF, and prior atrial ablation.

To define prerequisites for successful and safe discharge after AT ablation, the associations of baseline patient characteristics, procedural features, and early post-procedural events with LOS and subsequent clinical outcomes were evaluated using multivariable Cox models. Sensitivity analyses were conducted to determine the association between LOS and clinical outcomes by modelling LOS as a categorical, ordinal, and continuous variable, and by including interaction terms between LOS and LA procedures.

To explore system-level implications of discharge timing, we performed a model-based policy simulation using the day-2 landmark Cox models. Analyses were restricted to clinically stable patients without early post-procedural events. Follow-up time was reset to zero at day 2, and simulations were reported over the observed period of up to 180 days. Resource and cost implications were estimated by combining the simulated differences in clinical events with unit-cost assumptions (600€ per inpatient night, 3,000€ per unplanned cardiovascular hospitalization, and 8,000€ per repeat ablation) and are presented per 1,000 patients.^15–17^ For descriptive health services analyses, LOS was dichotomized into ≤1 vs. ≥2 nights to reflect commonly proposed early discharge strategies in routine clinical practice. All analyses were performed using GraphPad Prism 10.6.1 (GraphPad Inc., La Jolla, CA) and R 4.5.1 (R Foundation for Statistical Computing).

## Results

### Patient characteristics

A total of 670 consecutive patients undergoing AT catheter ablation were included. Median age was 67 [IQR 56–75] years and 54.9% were male. The most prevalent cardiac comorbidities were AF (66.7%), arterial hypertension (62.7%) and coronary artery disease (23.9%). Patients had a median CHA_2_DS_2_-VA score of 2 [IQR 1–3]. Prior atrial ablation was performed in 60.3%, previous cardiac surgery in 15.9% (Table 1). In our cohort, 8.1% of patients experienced acute minor complications and 12.5% showed early recurrences of atrial arrhythmia, resulting in 18.1% with a medically relevant event that could influence discharge timing.

**Table 1.** Baseline characteristics of the study population.

| Variable | Study population (n=670) | LOS ≤1 night (n=93) | LOS 2 nights (n=281) | LOS >2 nights (n=296) | P value |
| --- | --- | --- | --- | --- | --- |
| Age, years | 67 [56–75] | 67 [57–74] | 66 [55.5–73] | 68.5 [57–76] | 0.1528 |
| Male sex | 368 (54.9) | 64 (68.8)<sup>2,3</sup> | 153 (54.5)<sup>1</sup> | 151 (51.0)<sup>1</sup> | **0.0105** |
| Body mass index, kg/m<sup>2</sup> | 26.9±5.2 | 27.1±5.1 | 26.8±4.9 | 27.0±5.5 | 0.7749 |
| CHA<sub>2</sub>DS<sub>2</sub>-VA score | 2 [1–3] | 2 [1–3] | 2 [1–3] | 2 [1–3] | **0.0359** |
| Atrial fibrillation | 448 (66.7) | 53 (57.0)<sup>2</sup> | 194 (69.0)<sup>1</sup> | 201 (67.9) | 0.0890 |
| Arterial hypertension | 420 (62.7) | 55 (59.1) | 171 (60.9) | 194 (65.5) | 0.3802 |
| Coronary artery disease | 160 (23.9) | 18 (19.4) | 60 (21.4) | 82 (27.7) | 0.1100 |
| LVEF, % | 60 [50–60] | 60 [56.3–60]<sup>2,3</sup> | 60 [53–60]<sup>1</sup> | 57 [49–60]<sup>1</sup> | **0.0046** |
| Diabetes mellitus | 122 (18.2) | 20 (21.5) | 47 (16.7) | 55 (18.6) | 0.5709 |
| Prior stroke | 68 (10.1) | 12 (12.9) | 30 (10.7) | 26 (8.8) | 0.4809 |
| Vascular disorder | 99 (14.8) | 12 (12.9) | 47 (16.7) | 40 (13.5) | 0.4766 |
| Valvular disease | 127 (19.0) | 19 (20.4) | 60 (21.4) | 48 (16.2) | 0.2687 |
| Previous atrial ablation | 404 (60.3) | 37 (39.8)<sup>2,3</sup> | 176 (62.6)<sup>1</sup> | 191 (64.5)<sup>1</sup> | **<0.0001** |
| Duration since last ablation, months | 15 [6–39] | 16 [9–34] | 18 [6–42] | 14 [5–35] | 0.2961 |
| Previous cardiac surgery | 106 (15.8) | 14 (15.1) | 37 (13.2) | 55 (18.6) | 0.1999 |
| Antiarrhythmic drugs | 178 (26.6) | 15 (16.1)<sup>3</sup> | 65 (23.1)<sup>3</sup> | 98 (33.1)<sup>1,2</sup> | **0.0012** |
| Class I | 81 (12.1) | 10 (10.8) | 26 (9.3)<sup>3</sup> | 45 (15.2)<sup>2</sup> | 0.0828 |
| Amiodarone | 73 (10.9) | 4 (4.3)<sup>2,3</sup> | 32 (11.4)<sup>1</sup> | 37 (12.5)<sup>1</sup> | 0.0812 |
| Beta-receptor blockers | 445 (66.4) | 65 (69.9) | 181 (64.4) | 199 (67.2) | 0.5777 |
| Oral anticoagulation | 459 (68.5) | 67 (72.0) | 192 (68.3) | 200 (67.6) | 0.7174 |

Values are presented as mean ± SD, median [IQR] or n (%). LVEF = left ventricular ejection fraction. <sup>1</sup> = significant difference compared with group 1 (LOS ≤ 1 night), <sup>2</sup> = significant difference compared with group 2 (LOS 2 nights), <sup>3</sup> = significant difference compared with group 3 (LOS >2 nights).

### Patient and procedural characteristics according to hospitalization duration

LOS differed markedly according to patient and procedural characteristics (Table 1). Patients hospitalized for ≤1 night were more often male compared with those staying 2 or >2 nights (68.8% vs. 54.5% vs. 51.0%; *P*=0.0105), had a higher left ventricular ejection fraction (60% [56.3–60] vs. 60% [53–60] vs. 57% [49–60]; *P*=0.0046) and underwent prior atrial ablation less often (39.8% vs. 62.6% vs. 64.5%; *P*<0.0001). They presented less frequently with concomitant AF than patients hospitalized for 2 nights (57.0% vs. 69.0%; *P*=0.0334), without any difference compared with >2 nights, and were treated with amiodarone less often than those staying 2 (4.3% vs. 11.3%; *P*=0.0446) or >2 nights (4.3% vs. 12.5%; *P*=0.0247).

Procedure and fluoroscopy duration were shortest in patients discharged after ≤1 night, who also less frequently underwent LA ablation (26.5% vs. 76.0% vs. 80.8%; *P*<0.0001) (Figure 1). Additionally, patients with a LOS of 2 nights had a higher acute success rate than those staying >2 nights (97.9% vs. 94.6%; *P*=0.0403), with a comparable rate in those discharged after ≤1 night (Table 2).

**Figure 1.**
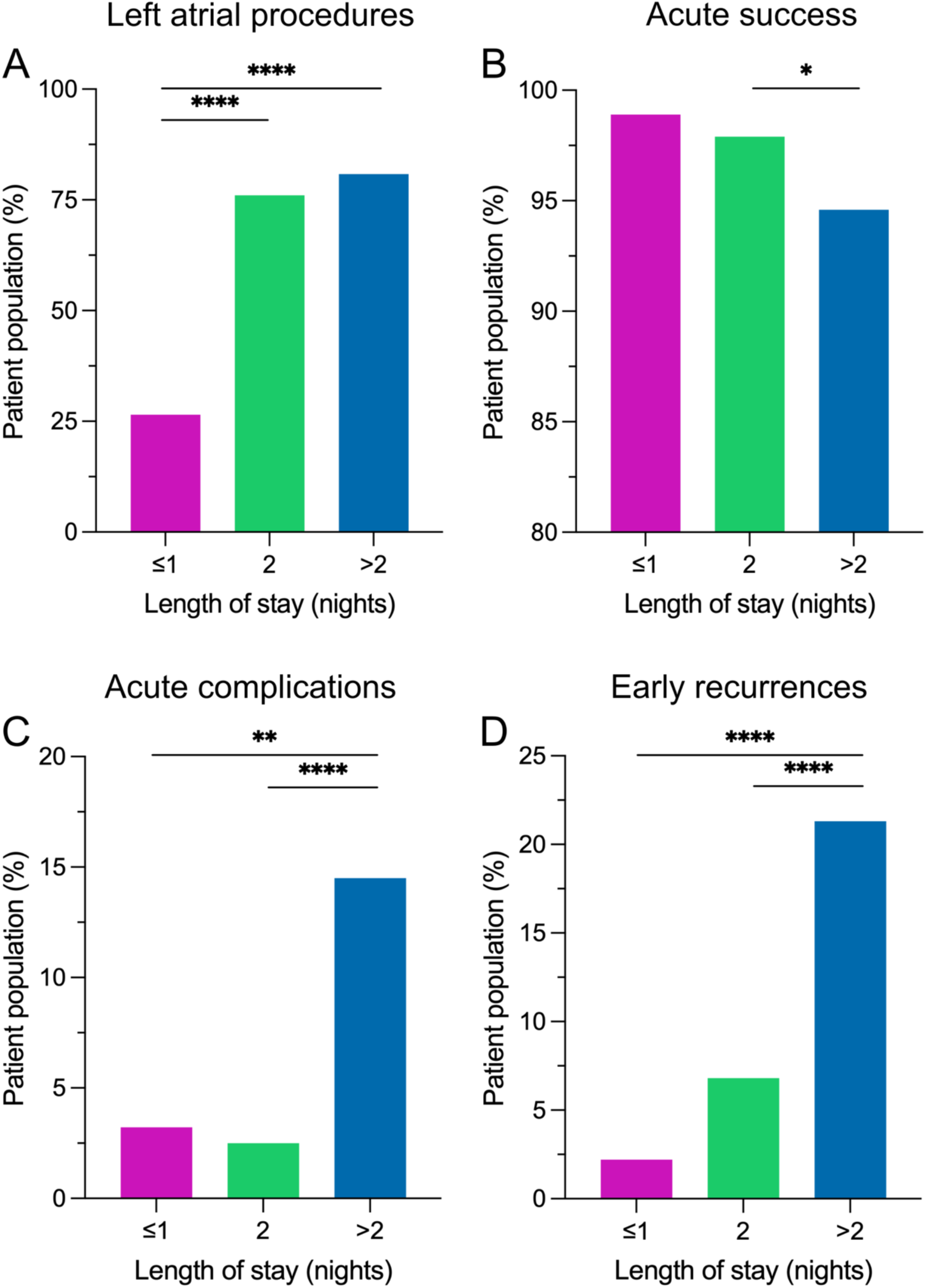
Determinants of hospitalization duration after AT ablation. A, Left atrial procedures, B, acute success, C, acute minor complications and D, early recurrences of atrial arrhythmia according to index length of stay are illustrated. **** *P*<0.0001; ** *P*<0.01; * *P*<0.05.

**Table 2.** Procedural characteristics of the study population.

| Variable | Study population (n=670) | LOS ≤1 night (n=93) | LOS 2 nights (n=281) | LOS >2 nights (n=296) | P value |
| --- | --- | --- | --- | --- | --- |
| Procedure duration, min | 164.7±71.6 | 123.1±64.3<sup>2,3</sup> | 169.0±62.1<sup>1</sup> | 173.4±77.1<sup>1</sup> | **<0.0001** |
| Fluoroscopy duration, min | 14.6±11.0 | 10.3±7.7<sup>2,3</sup> | 14.4±9.7<sup>1</sup> | 16.0±12.5<sup>1</sup> | **<0.0001** |
| AT cycle length, ms | 290 [250–370] | 295 [244–385] | 290 [250–350] | 305 [250–385]<sup>2</sup> | 0.3667 |
| Acute procedural success | 647 (96.6) | 92 (98.9) | 275 (97.9)<sup>3</sup> | 280 (94.6)<sup>2</sup> | **0.0395** |
| Early recurrence of atrial arrhythmia | 84 (12.5) | 2 (2.2)<sup>3</sup> | 19 (6.8)<sup>3</sup> | 63 (21.3)<sup>1,2</sup> | **<0.0001** |
| Acute complication | 53 (7.9) | 3 (3.2)<sup>3</sup> | 7 (2.5)<sup>3</sup> | 43 (14.5)<sup>1,2</sup> | **<0.0001** |
| Minor groin complication | 12 (1.8) | 1 (1.1) | 4 (1.4) | 7 (2.4) | 0.5943 |
| Major groin complication requiring intervention | 9 (1.3) | 0 (0) | 1 (0.4)<sup>3</sup> | 8 (2.7)<sup>2</sup> | **0.0240** |
| Vascular complication | 1 (0.1) | 0 (0) | 0 (0) | 1 (0.3) | 0.5312 |
| Respiratory insufficiency | 1 (0.1) | 0 (0) | 0 (0) | 1 (0.3) | 0.5312 |
| Pericardial effusion not requiring pericardiocentesis | 14 (2.1) | 1 (1.1) | 1 (0.4)<sup>3</sup> | 12 (4.1)<sup>2</sup> | **0.0062** |
| Pericardial effusion requiring pericardiocentesis | 8 (1.2) | 0 (0) | 0 (0)<sup>3</sup> | 8 (2.7)<sup>2</sup> | **0.0060** |
| Atrioventricular block requiring pacemaker implantation | 3 (0.4) | 0 (0) | 0 (0) | 3 (1.0) | 0.1490 |
| Cerebral embolic event with complete clinical recovery | 5 (0.7) | 1 (1.1) | 1 (0.4) | 3 (1.0) | 0.6067 |

Values are presented as mean ± SD, median [IQR] or n (%). <sup>1</sup> = significant difference compared with group 1 (LOS ≤1 night), <sup>2</sup> = significant difference compared
3 with group 2 (LOS 2 nights), <sup>3</sup> = significant difference compared with group 3 (LOS >2 nights).

### Determinants of hospitalization duration

Acute complications (3.2% vs. 2.5% vs. 14.5%; *P*<0.0001) resulted in longer LOS, largely driven by a higher rate of major groin complications requiring intervention and pericardial effusion in patients hospitalized for >2 compared with 2 nights (Figure 1). Similarly, early recurrences of atrial arrhythmia (2.2% vs. 6.8% vs. 21.3%; *P*<0.0001) led to longer LOS, with less frequent spontaneous termination in patients with a LOS of >2 vs. 2 nights (18.2% vs. 62.5%; *P*=0.0005). Clinical and procedural predictors of acute minor complications and early recurrences of atrial arrhythmia and the composite of both are listed in Table 3.

**Table 3.** Predictors of acute minor complications and early recurrences of atrial arrhythmia after AT ablation.

| Variable | Acute minor complication |  | Early recurrence of atrial arrhythmia |  | Composite endpoint |  |
| --- | --- | --- | --- | --- | --- | --- |
|  | OR (95% CI) | *P* value | OR (95% CI) | *P* value | OR (95% CI) | *P* value |
| Age, per years | 0.99 [0.96–1.03] | 0.7299 | 0.99 [0.96–1.02] | 0.4182 | 0.99 [0.97–1.02] | 0.8824 |
| Male sex | 0.77 [0.37–1.60] | 0.4875 | 0.81 [0.44–1.49] | 0.5054 | 0.93 [0.55–1.59] | 0.7924 |
| CHA<sub>2</sub>DS<sub>2</sub>-VA score | 1.34 [0.95–1.89] | 0.0987 | 1.20 [0.90–1.59] | 0.2126 | 1.13 [0.88–1.45] | 0.3408 |
| Atrial fibrillation | 0.34 [0.11–1.04] | 0.0582 | 1.01 [0.42–2.51] | 0.9760 | 0.66 [0.30–1.45] | 0.2998 |
| Prior atrial ablation | 1.03 [0.38–3.00] | 0.9547 | 0.49 [0.22–1.13] | 0.0924 | 0.61 [0.29–1.30] | 0.19555 |
| Procedure duration, per 30 min | 1.27 [1.11–1.47] | **0.0006** | 1.12 [0.98–1.27] | 0.0896 | 1.22 [1.10–1.37] | **0.0003** |
| Left atrial ablation | 1.04 [0.44–2.64] | 0.9369 | 1.27 [0.62–2.73] | 0.5191 | 1.13 [0.60–2.21] | 0.7004 |
| ≥1 reentrant AT | 3.17 [1.18–9.78] | **0.0206** | 1.96 [0.89–4.67] | 0.0964 | 2.90 [1.41–6.44] | **0.0033** |
| Acute procedural success | - | - | 0.13 [0.04–0.41] | **0.0007** | 0.13 [0.04–0.38] | **0.0003** |

Values are odds ratios (ORs) with 95% CIs derived from Firth penalized multivariable logistic regression.

In a clinical model examining predictability of an additional hospital night (≤1 vs. 2 nights), higher age (OR 0.96, 95% CI 0.94–0.99; *P*=0.0014) and male sex (OR 0.48, 95% CI 0.29–0.82; *P*=0.0071) were independently associated with a lower likelihood of a 2-night stay, whereas concomitant AF (OR 1.91, 95% CI 1.06–3.42; *P*=0.0302) and prior atrial ablation (OR 2.18, 95% CI 1.27–3.75; *P*=0.0048) increased the probability (Table 4). Overall discrimination of the clinical model was modest (AUC 0.73), suggesting limited ability to anticipate discharge timing before the early post-procedural course. The addition of procedural variables resulted in a numerical improvement in discrimination (AUC 0.76), and LA ablation was associated with higher odds of a 2-night hospital stay (OR 3.0, 95% CI 1.19–7.54; *P*=0.0197). Incorporation of early post-procedural events further improved discrimination (AUC 0.77), with a significant increase compared with the procedural model (ΔAUC +0.013; DeLong *P*=0.0300) (Figure 2A). Consistently, when comparing ≤1 vs. ≥2 nights, the clinical model demonstrated modest discrimination (AUC 0.72). While addition of procedural variables resulted in a numerical increase in discrimination (AUC 0.76), inclusion of early post-procedural events significantly improved discrimination (AUC 0.80), both compared with the clinical model (ΔAUC +0.074; DeLong *P*=0.0063) and the procedural model (ΔAUC +0.041; DeLong *P*=0.0003). These findings indicate that LOS after AT ablation is primarily driven by the immediate peri-procedural course and only modestly predictable from baseline clinical and procedural factors, suggesting that LOS captures clinically relevant medical pathways rather than purely administrative prolongation. When patients were stratified into tertiles of the clinical linear predictor, observed event rates of LOS ≥2 nights increased from 75.9% in the low-risk group to 89.7% and 92.8% in the intermediate- and high-risk group. Calibration across deciles demonstrated acceptable agreement between predicted probabilities (66.1%–96.3%) and observed event rates (58.2%–94.0%), albeit with limited risk separation in a cohort with a generally high prevalence of LOS ≥2 nights (Figure 2B+C).

**Figure 2.**
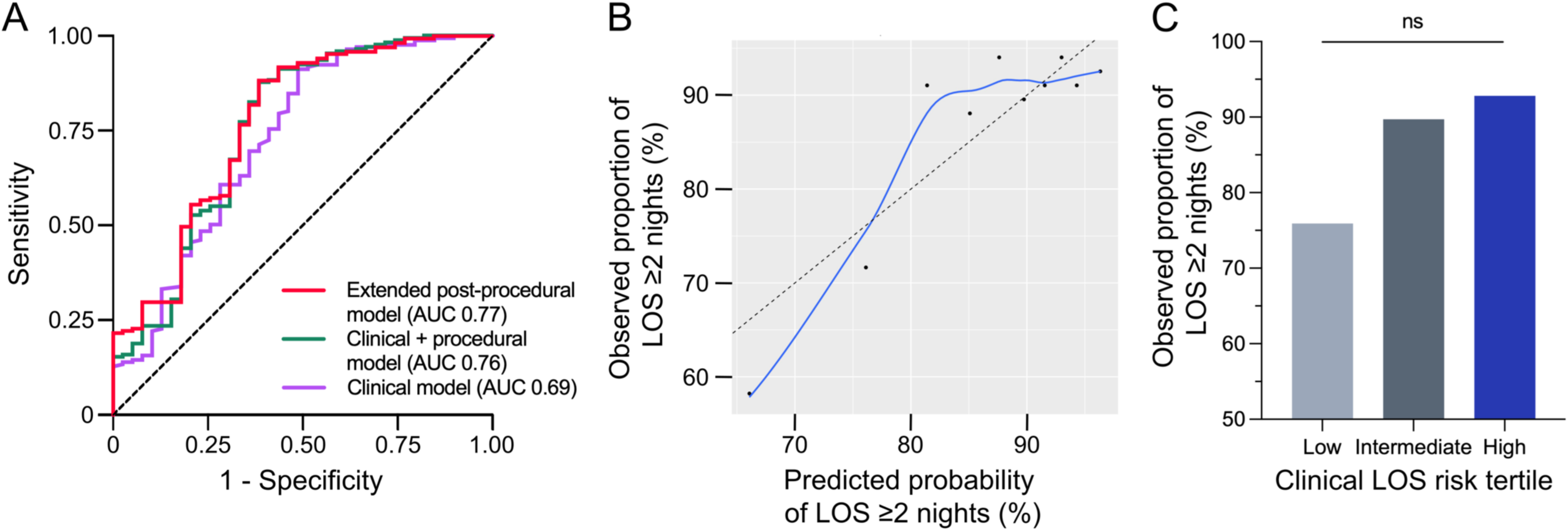
Discriminative performance of hierarchical models for prediction of hospitalization duration after AT ablation. A, Receiver operating characteristic curves illustrating the discriminative performance of hierarchical logistic regression models for prediction of an additional hospital night after AT ablation (≤1 vs. 2 nights). Model 1 included baseline clinical characteristics only, model 2 additionally incorporated procedural variables, and model 3 further included early post-procedural events. Discrimination was quantified using the area under the curve (AUC). B, Observed proportions of LOS ≥2 nights are plotted against mean predicted probabilities across deciles of predicted risk. The LOESS fit illustrates overall calibration, with a plateau at high predicted risk reflecting the high prevalence of prolonged LOS in the cohort. C, Distribution of patients across the three tertiles of clinical LOS risk, with the bar plot showing the percentage of patients within each tertile (low, intermediate, high) who experienced LOS ≥2 nights. The results illustrate the increasing likelihood of prolonged hospitalization as clinical risk increases, with the high tertile exhibiting the highest percentage of patients requiring longer stays. However, no significant difference was observed between the tertiles, indicating the overall high proportion of prolonged LOS.

**Table 4.** Multivariable models for prediction of hospitalization duration after AT ablation.

| Variable | Model 1: Clinical |  | Model 2: Clinical + procedural |  | Model 3: Extended post-procedural |  |
| --- | --- | --- | --- | --- | --- | --- |
|  | OR (95% CI) | *P* value | OR (95% CI) | *P* value | OR (95% CI) | *P* value |
| Age, per years | 0.96 [0.94–0.98] | **0.0014** | 0.97 [0.93–1.01] | 0.1596 | 0.97 [0.93–1.01] | 0.1900 |
| Male sex | 0.48 [0.29–0.82] | **0.0071** | 0.45 [0.19–1.06] | 0.0677 | 0.45 [0.19–1.07] | 0.0721 |
| CHA<sub>2</sub>DS<sub>2</sub>-VA score | 1.21 [0.96–1.53] | 0.1140 | 1.04 [0.71–1.53] | 0.8430 | 1.05 [0.71–1.55] | 0.8230 |
| Atrial fibrillation | 1.91 [1.06–3.42] | **0.0302** | 1.53 [0.47–5.01] | 0.4831 | 1.51 [0.46–5.01] | 0.4970 |
| Prior atrial ablation | 2.18 [1.27–3.75] | **0.0048** | 3.21 [1.17–8.82] | **0.0235** | 3.41 [1.23–9.44] | **0.0180** |
| Procedure duration, per 30 min | - | - | 1.08 [0.89–1.32] | 0.4208 | 1.08 [0.89–1.33] | 0.4510 |
| Left atrial ablation | - | - | 3.00 [1.19–7.54] | **0.0197** | 2.96 [1.18–7.43] | **0.0208** |
| ≥1 reentrant AT | - | - | 0.49 [0.17–1.42] | 0.1901 | 0.46 [0.16–1.35] | 0.1570 |
| Acute procedural success | - | - | 3.84 [0.19–79.74] | 0.3842 | 4.13 [0.16–109.0] | 0.3960 |
| Acute minor complication | - | - | - | - | 1.24 [0.06–24.80] | 0.8870 |
| Early recurrence of atrial arrhythmia | - | - | - | - | 3.56 [0.36–35.0] | 0.2760 |
| **Discrimination** | **0.729** |  | **0.760** |  | **0.773** |  |
| **DeLong *P* for ΔAUC** | - |  | M1 vs. M2: 0.2196 |  | M1 vs. M3: 0.0830<br>**M2 vs. M3: 0.0300** |  |

3 Multivariable models comparing discharge after ≤1 vs. 2 nights. Values are presented as ORs with 95% CIs. Model discrimination was quantified using the area 4 under the receiver operating characteristic curve (AUC). Incremental discrimination between nested models was assessed using DeLong's test based on refitted 5 complete-case models.

### Clinical outcomes according to hospitalization duration

During a follow-up of 2.8±3.0 years, subsequent cardiovascular hospitalization occurred numerically less often among patients discharged after ≤1 night compared with 2 or >2 nights (41.9% vs. 58.4% vs. 58.4%; *P*=0.0189), although time to first cardiovascular hospitalization did not differ (Figure 3). Repeat ablation was more frequent in patients with a LOS of 2 vs. >2 nights (39.1% vs. 31.1%; *P*=0.0424), whereas index AT mechanism (non-peritricuspid macro-reentrant AT in 71.7% vs. 76.8%; *P*=0.1997), arrhythmia recurrences (47.3% vs. 45.9%; *P*=0.7684) and time to first repeat ablation were comparable (Figure 4). Notably, patients discharged after ≤1 or 2 nights more commonly underwent only a single repeat ablation compared with those hospitalized for >2 nights (78.6% vs. 80.9% vs. 62.4%; *P*=0.0096).

**Figure 3.**
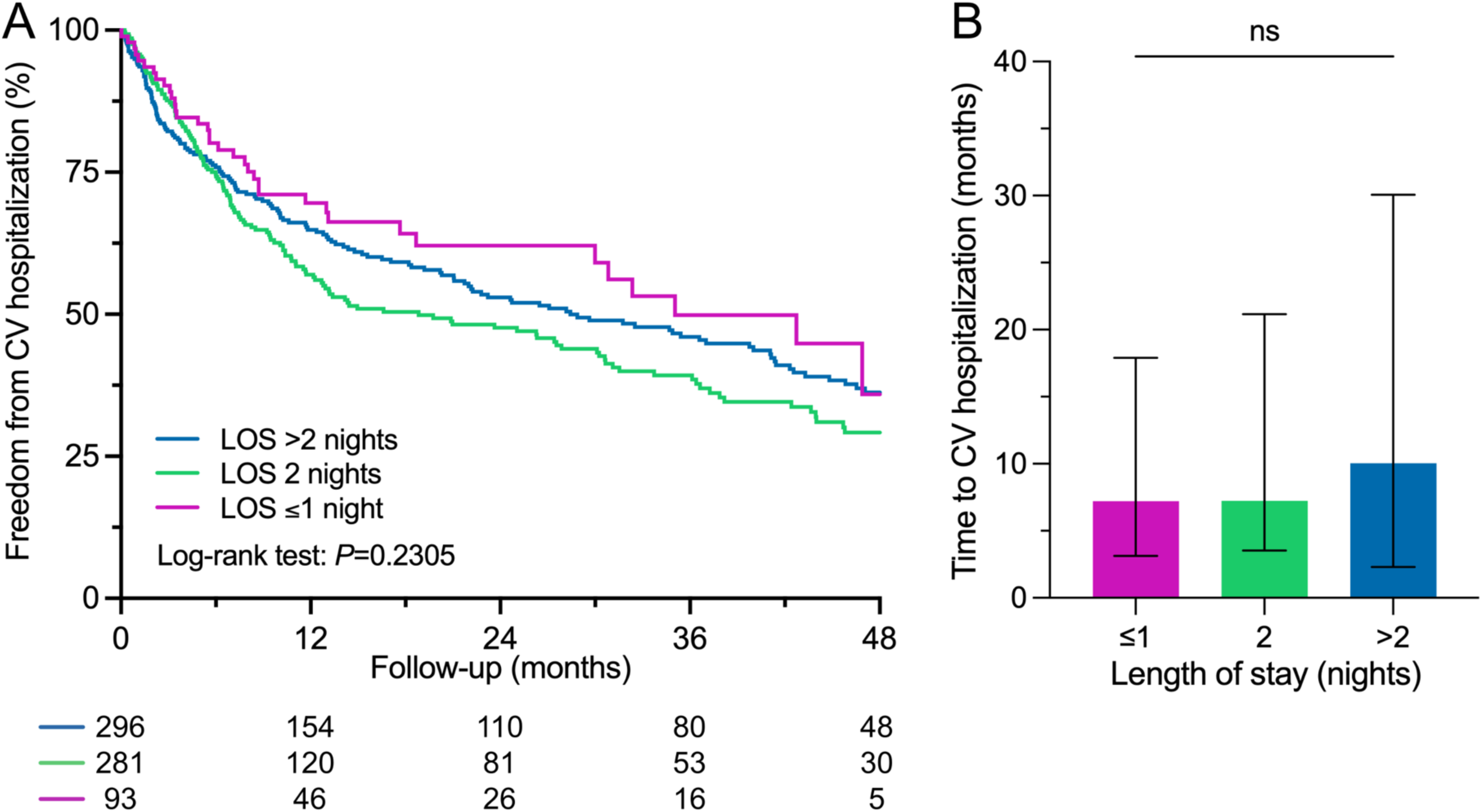
Freedom from CV hospitalization according to hospitalization duration after AT ablation. A, Freedom from CV hospitalization and B, time to first CV hospitalization according to index LOS are illustrated. CV, cardiovascular; LOS, length of stay; ns, not significant.

**Figure 4.**
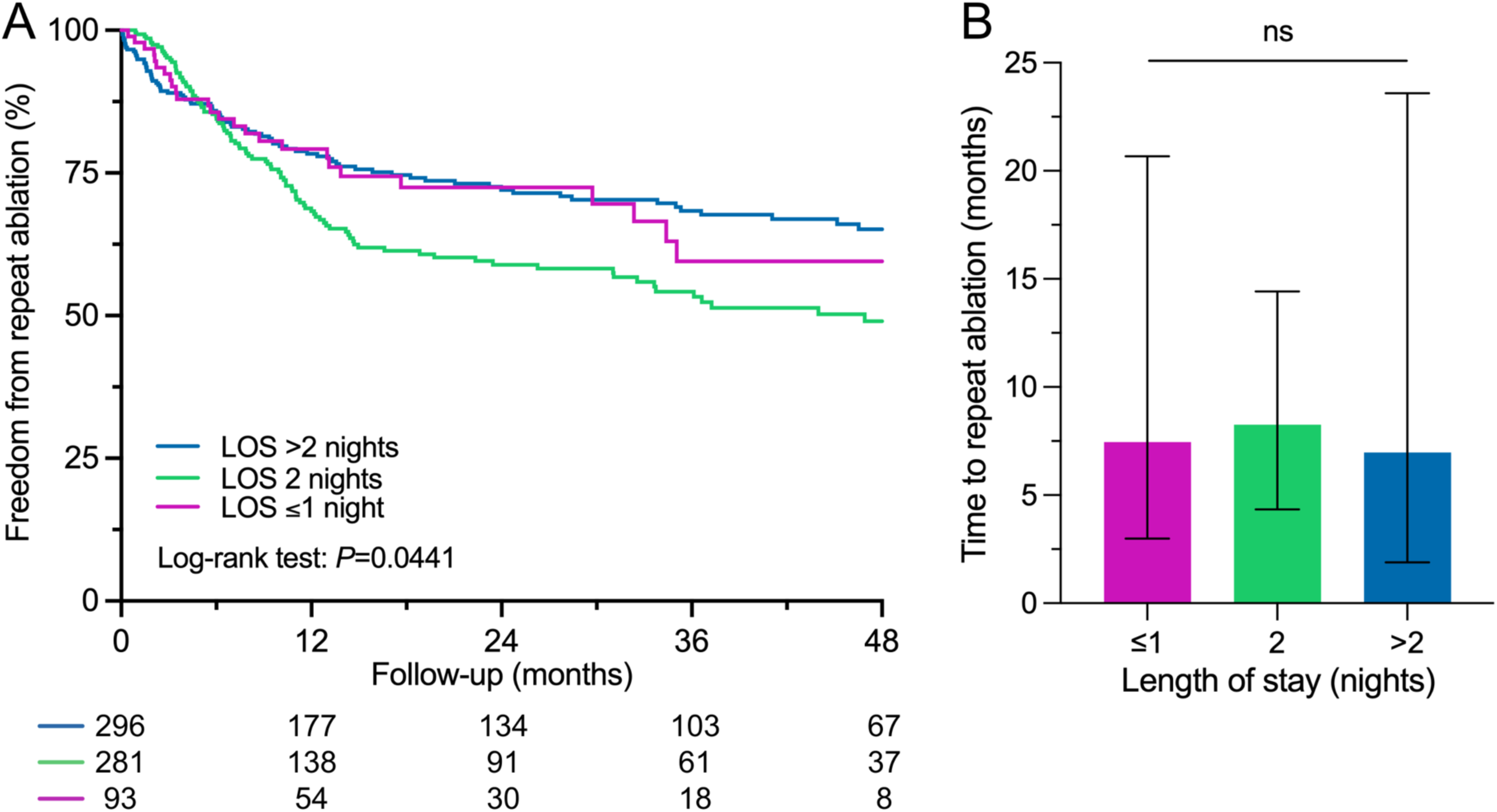
Freedom from repeat ablation according to hospitalization duration after AT ablation. A, Freedom from repeat ablation and B, time to first repeat ablation according to index LOS are illustrated. LOS, length of stay; ns, not significant.

In a landmark analysis at day 2 restricted to clinically stable patients without early post-procedural events and discharged after ≤1 or 2 nights, LOS was not associated with subsequent cardiovascular hospitalization (HR 1.19, 95% CI 0.78–1.81; *P*=0.4175) or repeat ablation (HR 1.11, 95% CI 0.69–1.81; *P*=0.6611). Instead, comorbidity burden emerged as the principal determinant of long-term outcomes. AF was independently associated with both hospitalization and repeat ablation, while a higher CHA₂DS₂-VA score predicted hospitalization and increasing age was inversely associated with repeat ablation (Figure 5). These findings indicate that among clinically stable patients, extending hospitalization beyond one night does not provide additional prognostic benefit, whereas patient-related factors remain the main drivers of adverse events during long-term follow-up.

**Figure 5.**
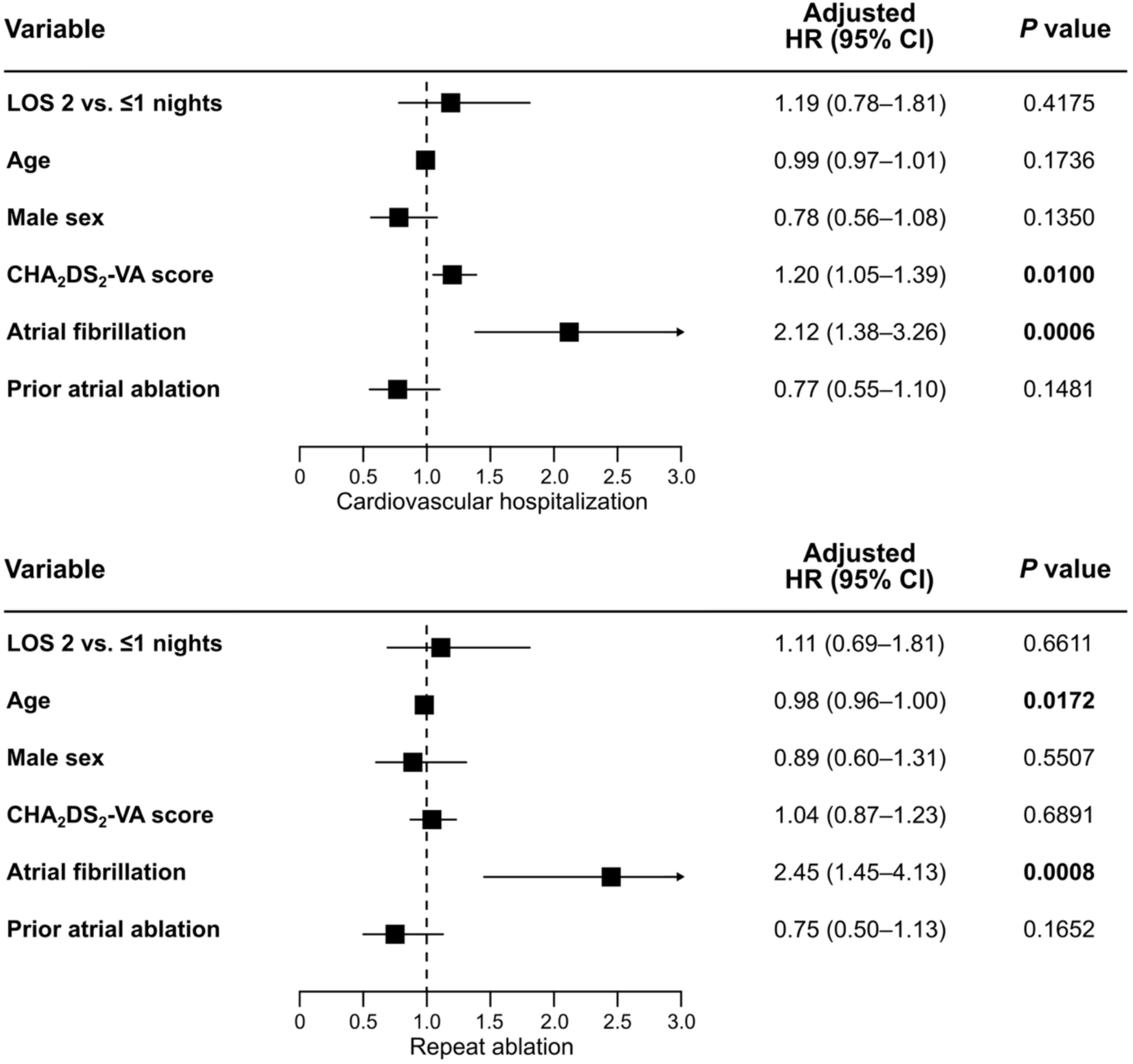
Landmark-adjusted predictors of clinical outcomes after AT ablation. Multivariable Cox proportional hazards models with a landmark set at day 2. Analyses were restricted to patients without acute minor complications or early recurrences of atrial arrhythmia. Results are reported as hazard ratios (HRs) with 95% confidence intervals (CIs) and *P* values.

### Prognostic value of early post-procedural events

In adjusted Cox regression analyses, early recurrences of atrial arrhythmia were associated with an increased risk of repeat ablation during long-term follow-up (HR 1.89, 95% CI 1.34–2.67; *P*=0.0003), which was confirmed in a landmark analysis at day 2 restricted to patients without acute minor complications (HR 1.80, 95% CI 1.23–2.65; *P*=0.0027), underscoring the prognostic relevance of early post-procedural rhythm instability. In contrast, acute minor complications were not independently linked with cardiovascular hospitalization (HR 1.04, 95% CI 0.73–1.49; *P*=0.8300) or repeat ablation (HR 0.98, 95% CI 0.62–1.56; *P*=0.9410).

### Sensitivity and robustness analyses

In multivariable Cox regression analyses adjusting for clinical covariates, LOS was not associated with cardiovascular hospitalization or repeat ablation. In contrast, patient-related factors reflecting comorbidity burden were the primary determinants of clinical outcomes (Table 5). When early recurrence of atrial arrhythmia was included in the multivariable models, LOS again was not associated with cardiovascular hospitalization (2 vs. ≤1 night: HR 1.25, 95% CI 0.87–1.81; *P*=0.2294; >2 vs. ≤1 night: HR 0.94, 95% CI 0.64–1.38; *P*=0.7525). Instead, early recurrence of atrial arrhythmia was an independent predictor of hospitalization (HR 1.48, 95% CI 1.07–2.05; *P*=0.0189), alongside AF and a higher CHA₂DS₂-VA score. Sensitivity analyses modelling LOS as a categorical, ordinal, or continuous variable using the same hierarchical covariate structure including procedural factors and early post-procedural events yielded consistent results, supporting LOS as a marker of early post-procedural risk rather than a causal determinant of long-term outcomes. In the fully adjusted ordinal model, acute minor complications remained strongly associated with longer LOS (OR 2.92, 95% CI 1.42–6.44; *P*=0.0033), as did early recurrences of atrial arrhythmia (OR 2.90, 95% CI 1.41–6.44; *P*=0.0028), whereas baseline characteristics were attenuated and no longer statistically significant.

**Table 5.**
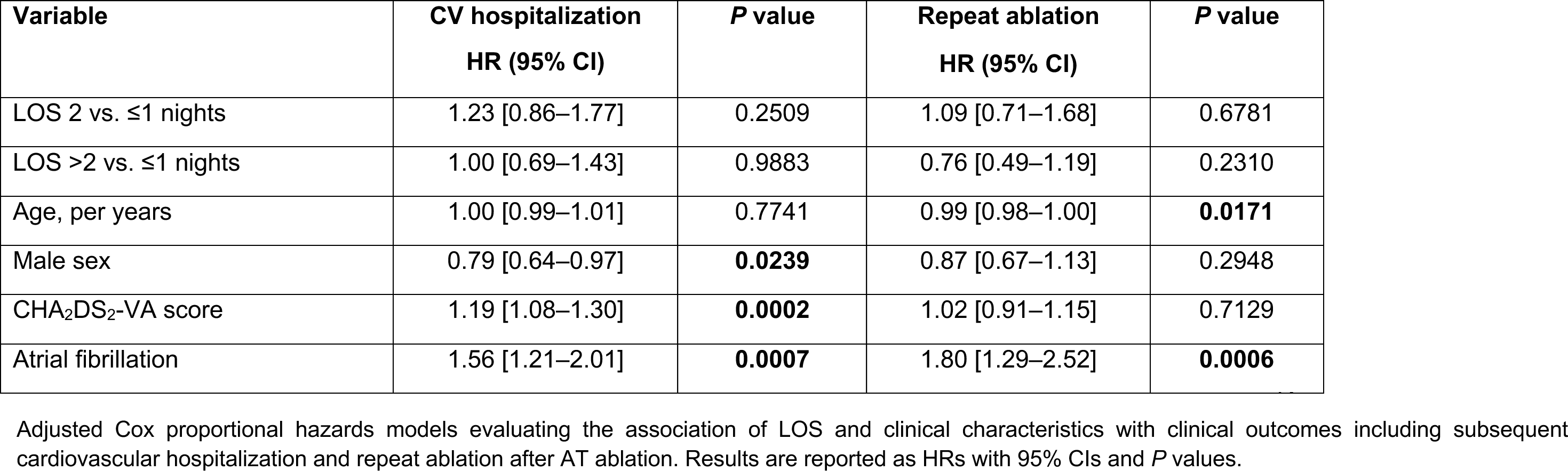
Adjusted Cox regression models for clinical outcomes after AT ablation.

In multivariable logistic regression analyses, LA procedures were a strong independent determinant of prolonged hospitalization with a more than fivefold higher odds of a LOS of ≥2 nights (OR 5.55, 95% CI 3.07–10.05; *P*<0.0001). In Cox proportional hazards models including LOS category, LA procedures, and their interaction, LOS was not independently associated with cardiovascular hospitalization or repeat ablation, and interaction terms were not significant (hospitalization: *P* for interaction = 0.7806; repeat ablation: *P* for interaction = 0.1767). Accordingly, LA procedures were associated with longer hospitalization but did not modify the lack of prognostic relevance of LOS for long-term clinical outcomes.

### Policy simulation of discharge strategies

In a policy simulation, discharge after ≤1 night in clinically stable patients without early post-procedural events was not associated with increased healthcare utilization within 180 days after the landmark. Compared with patients needing 2 nights of hospitalization, earlier discharge was associated with 16 fewer cardiovascular hospitalizations and 28 fewer repeat ablations per 1,000 patients. These findings suggest that earlier discharge in clinically stable patients undergoing AT ablation may not be associated with harm. Because 2-night stays were common in the present cohort, an early discharge approach resulted in an estimated reduction of 736 inpatient bed-days per 1,000 patients. Based on the applied unit-cost assumptions, this corresponded to an estimated net cost saving of approximately €0.71 million per 1,000 patients within 180 days.

In routine clinical practice, hospitalization for ≥2 nights occurred in 861 per 1,000 patients, indicating that adoption of a strict 1-night discharge standard would have necessitated extension of stay in the majority of cases. From a health care services perspective, such extensions imply substantial inpatient resource utilization and underscore that prolonged hospitalization after AT ablation primarily reflects peri-procedural monitoring needs within routine clinical practice, thereby challenging rigid, logistics-driven discharge targets.

## Discussion

In this multicenter study, we analyzed for the first time patient- and procedure-related prerequisites for successful and safe discharge strategies after AT catheter ablation. In summary, we found that 1) despite a high comorbidity burden, most patients undergoing AT ablation need up to 2 nights of hospitalization, 2) prolonged hospital stays before successful and safe discharge are common, associated with acute minor complications and early recurrences of atrial arrhythmia rather than comorbidities and therefore difficult to predict and 3) among clinically stable patients, extended hospitalization does not improve outcomes.

### Hospitalization duration after AT ablation

In the present study, most patients needed up to 2 nights of hospitalization after AT ablation, which is notable given the advanced age and substantial comorbidity burden of the cohort. At the same time, marked heterogeneity in baseline characteristics was observed. Patients discharged after ≤1 night consistently exhibited a more favourable clinical profile, resembling selected AF ablation populations in whom early or even same-day discharge has been shown to be feasible and safe without increased adverse events or rehospitalizations.^6^ This is in line with contemporary AF ablation data showing a temporal decline in procedure-related complications, largely attributed to advances in ablation technologies and shorter procedure durations.^18^ However, AT ablation differs from AF ablation in several aspects directly relevant to discharge planning.^12^ Procedures more frequently target complex, scar-related or post-surgical substrates,^19^ are characterized by greater uncertainty due to inducibility and mapping dependence and are followed by early recurrences of atrial arrhythmia in a substantial proportion of patients.^2,10^ These mechanistic differences indicate that early discharge strategies established for AF ablation cannot be directly applied to AT. Accordingly, the frequent need for prolonged hospitalization observed in our cohort, particularly among patients undergoing LA ablation and longer procedures, underscores the importance of individualized post-procedural monitoring in AT patients, despite ongoing technical advances.

### Determinants of hospitalization duration

The present findings indicate that hospitalization duration after AT ablation is only modestly predictable from clinical characteristics alone. Although factors such as age, sex, concomitant AF, and prior ablation were associated with LOS, their ability to guide discharge planning in advance was limited. In contrast to AF ablation, where pre-procedural risk stratification has been more successfully used to support early discharge strategies,^4,5^ procedural complexity, particularly LA ablation, was associated with a higher likelihood of prolonged hospitalization, reflecting increased monitoring needs. Importantly, a second night may in some patients primarily reflect the need for clinical observation or telemetry monitoring, whereas in others additional medical or organizational considerations and diagnosis-related group structures may contribute, underscoring that LOS represents a mixed endpoint rather than a direct surrogate of medical necessity. However, the strongest determinants of prolonged LOS were early post-procedural events. Patients with longer hospital stays showed a markedly higher rate of acute minor complications and early recurrences of atrial arrhythmia, which were less likely to terminate spontaneously and often necessitated continued in-hospital observation. Together, these findings indicate that prerequisites for early discharge after AT ablation extend beyond baseline patient and procedural characteristics and are primarily defined by the early post-procedural clinical course. This explains why discharge timing in this population is difficult to standardize and why flexible, clinically guided discharge pathways are preferable to fixed, pre-planned strategies. Early discharge concepts for AT ablation should therefore incorporate structured post-procedural monitoring rather than rely exclusively on pre-procedural stratification.

### Clinical implications of hospitalization duration

Despite its association with comorbidity burden and early post-procedural events, LOS itself did not independently predict long-term outcomes once patients were clinically stable. Importantly, early recurrences of atrial arrhythmia, but not acute minor complications, emerged as an independent predictor of clinical outcomes. This observation aligns with AF populations demonstrating that early arrhythmia recurrences reflect persistent arrhythmogenic substrate and predict late recurrences.^10,20^ In contrast, acute minor complications appeared to affect LOS without translating into adverse outcomes. In line with a previously proposed conceptual framework for outcome assessment in cardiovascular care,^21^ this finding therefore reinforces the concept that LOS primarily reflects early post-procedural instability rather than a modifiable determinant of prognosis.

Beyond procedural safety, psychological outcomes may be particularly relevant when considering early discharge after AT ablation. Evidence from AF ablation and other elective cardiac interventions^22,23^ suggests that early discharge is usually well accepted but may be associated with transient anxiety or uncertainty in a subset of patients, especially in the presence of post-procedural symptoms. However, data on psychological outcomes after early discharge in AT patients are currently lacking. This gap is clinically relevant, as early recurrences of atrial arrhythmia after AT ablation may occur and may potentially amplify patient concern after discharge. Future studies should therefore incorporate AT-specific patient-reported outcome measures to better inform individualized discharge strategies.

### Health care services and policy implications

From a health care services perspective, the observed LOS distribution highlights the limitations of applying uniform ambulatory care models to the heterogeneous AT population. While selected patients, particularly those with less complex atrial substrates and uncomplicated post-procedural courses, may be suitable for early discharge, a substantial proportion needs prolonged hospitalization. In this respect, complex AT ablation^24,25^ shares important similarities with ventricular tachycardia ablation, which is widely recognized as a technically challenging procedure associated with advanced structural heart disease.^26–28^ Since 2026, in the German healthcare system, left ventricular tachycardia ablation continues to represent the only electrophysiological procedure consistently reimbursed as inpatient care, reflecting its perceived risk profile and complexity. Our here presented policy simulation further supports this differentiated approach. Among clinically stable patients, discharge after ≤1 night was not associated with increased healthcare utilization but was projected to reduce hospitalizations, repeat procedures, and inpatient bed-days without evidence of harm. However, these patients had fewer comorbidities, which may have contributed to more favourable long-term outcomes. It is also important to note that prolonged hospital stays were justified by significant clinical surveillance needs, which, fortunately, did not result in additional effort or costs during follow-up. Although these estimates are exploratory and not causal, they support the notion that structured early discharge approaches may be considered in carefully selected AT patients. At the same time, flexibility to prolong hospitalization in the presence of early post-procedural events remains essential, while the majority of patients undergoing AT ablation will likely continue to need more than one night of hospitalization.

### Limitations

Several limitations merit consideration. First, discharge decisions were not randomized and may have been influenced by unmeasured clinical factors, reimbursement characteristics, patient preferences or center-specific practices. Second, the policy simulations are model-based projections rather than causal estimates and should be interpreted as informing population-level discharge strategies. Third, cost estimates were derived from unit-cost assumptions instead of patient-level billing data and may not fully capture indirect costs or inter-system variability. Finally, this analysis was conducted within the German healthcare system, where reimbursement structures and inpatient care pathways differ from those in other settings, which may limit the generalizability of these findings.

## Conclusion

Despite multiple comorbidities, most patients undergoing AT ablation need up to 2 nights of hospitalization. However, prolonged hospital stays before successful and safe discharge are common and associated with acute minor complications and early recurrences of atrial arrhythmia rather than comorbidities. Accordingly, discharge timing largely reflects the immediate peri-procedural clinical course, therefore challenging purely logistics-driven planning.

## Consent

The study was conducted in accordance with the provisions of the Declaration of Helsinki. Data collection and analysis were performed under a protocol approved by the institutional review boards. Patients gave written informed consent prior to all procedures.

## Author contributions

A.K.K. and C.M. designed the study; A.K.K., F.D., P.F., O.A., A.H., J.J., J.M. and K.A. collected the data; A.K.K. analyzed and interpreted the data and wrote the initial draft; C.M. supervised the study, managed project administration, interpreted the data, reviewed and edited the manuscript. All authors contributed to data interpretation, critically revised the manuscript, and approved the final version of the manuscript.

## Acknowledgements

Part of the Structured Graphical Abstract was created with BioRender.com.

## Data Availability

The data underlying this article will be shared on reasonable request to the corresponding author.

